# Predictors of breast cancer screening among women of reproductive age in Tanzania: Evidence from DHS 2022

**DOI:** 10.1101/2024.02.05.24302367

**Authors:** Jovin R. Tibenderana, Sanun Ally Kessy, Dosanto Felix Mlaponi, Ndinagwe Lloyd Mwaitete, John Elyas Mtenga

## Abstract

**Background:** Breast cancer is a global concern, registering 2.3 million new cases and 685,000 deaths in 2020, with projections reaching 4.4 million cases by 2070. In Tanzania, it’s the second leading cause of cancer-related deaths among women, often diagnosed at advanced stages, leading to poor outcomes. Only 5% of women in the country report undergoing breast cancer screening, the aim study is to determine factors associated with breast cancer screening in Tanzania.

**Methods:** A cross-sectional study among women of reproductive age in Tanzania, utilizing data from the Demographic and Health Surveys (DHS). We used available data on breast cancer screening the DHS. The outcome of the study was breast cancer screening. To find independent variables associated with breast cancer screening, logistic regression was used.

**Results:** After controlling for other factors, the following factors remained significantly associated with breast cancer screening among women of reproductive age; age(AOR=5.33, 95% CI 3.72, 7.63), being wealthy (AOR=2.34, 95% CI 1.61, 3.38), residing in rural(AOR= 0.59, 95% CI 0.46, 0.763), being educated(AOR=2.43, 95% CI 1.60, 3.68), being insured(AOR= 2.40, 95% CI 1.89, 3.06), healthcare facility visits in the past 12 months(AOR=1.43, 95% CI 1.14, 1.78) and living in Northern zone(AOR= 2.43, 95% CI 1.42, 4.15) compared to western zone

**Conclusion:** Breast cancer screening is still under-utilized and have shown to be marginalized in women of reproductive age. Policies to address disparities, comprehensive health education and awareness campaigns are instrumental to increase utilization and reduction of burden of breast cancers in Tanzania

## Introduction

In the year 2020, there were around 2.3 million newly diagnosed cases of breast cancer and 685,000 deaths attributed to breast cancer globally(1,2) and cases are expected to rise up to 4.4 million in 2070 (3). In the women population, breast cancer comprised about 24.5% of total cancer cases and 15.5% of cancer-related deaths, securing the top position in terms of both incidence and mortality in the majority of countries worldwide in 2020(2).

Low- and middle-income countries (LMICs) and developing countries have seen a sharp increase in the incidence and mortality rates of breast cancer. These countries also have much lower 5-year survival rates for breast cancer, at around 53%.(2,4,5). In Africa the focus has mostly been in communicable diseases like Tuberculosis, leading to less engagement in NCDs e.g. Cancers and hence very low rate of breast cancer screening(5).

After cervical cancer, breast cancer stands as the second most prevalent cancer and the second leading cause of cancer-related deaths among women in Tanzania(6). Tanzanian women are expected to have a lifetime risk of 1 in 203 of acquiring breast cancer, and more than 50% of those who receive a diagnosis will pass away from the disease and related complications.(7). In Tanzania 80% of all women diagnosed with breast cancer are diagnosed late at stage III and IV where outcome is poor and treatment is not effective (6–8). Notably awareness seems to still be a problem, Morse and colleagues reported that 44% never heard of Self Breast exam 32% never heard of Clinical breast examination(9).

In Tanzania, 14.4% of newly diagnosed malignancies in women are breast cancers. The number of newly diagnosed cases of breast cancer in Tanzania is expected to rise by 82% by 2030(6). Tanzania has an age-standardized incidence of 19.4/100,000 women with breast cancer and an age-standardized death rate of 9.7/100,000. This translates to a mortality-to-incidence ratio (MIR) of 0.5 (6,10,11). Among females 15.9% of all new diagnosed cancers in 2022 were breast cancer in Tanzania and ranked second for both males and females for the newly diagnosed cancers.(10).

Tanzania Demographic health survey 2022 has reported that, only 5% of women age 15–49 reported that they had been screened for breast cancer in Tanzania(12). Most of the studies in Tanzania have studied factors associated with awareness of breast cancer screening(6,9,13,14)

Screening can effectively reduce mortality, morbidity as well as poor quality of life from breast cancer(15,16). For early detection of breast cancer employment of methods like self-breast examination, clinical-breast examination and mammography must be employed. these methods are said to reduce the rate of mortality from breast cancer by 25-30%(17).

A multi-country study has revealed significant associations between breast cancer screening and various factors. These factors encompass higher educational attainment, advanced age, possession of health insurance, elevated socio-economic status, and ownership of a television(5). The true extent of breast cancer prevalence is not accurately represented in many Sub-Saharan African (SSA) countries, leading to underreporting and a lack of a genuine reflection of the disease burden(18,19).

Research on breast cancer screening in Tanzania are limited, little is known about factors associated with breast cancer screening in Tanzania(20). This baseline knowledge is essential in influencing educational programs that enhance comprehension and focus on evidence-based, lifestyle-oriented interventions for breast cancer screening and promoting early detection and treatment. It’s crucial therefore to identify factors affecting breast cancer screening, hence this study purpose is to determine factors associated with breast cancer screening in Tanzania.

## Material and Methods

### Study setting and period

The study utilized the Tanzania Demographic and Health Survey (TDHS) data, a nationwide cross-sectional survey conducted every 5 years (21). TDHS 2022 is the most recent data which collected data on breast cancer screening among women of reproductive age. Tanzania is the largest country in East Africa, covering 940,000 square kilometers, 60,000 of which are inland water. The population of Tanzania as of 2022 was estimated to be 61,741,120 with an annual population growth rate of 3.2% (21,22).

### Study design and data source

This was an analytical cross-sectional study conducted using nationally representative secondary data from the Tanzania demographic and health surveys (TDHS) of 2022. We explored women’s data. DHS is national representative data which is funded by the U.S Agency for International Development and implemented by the Ministry of Health (MoH) (Tanzania Mainland), Ministry of Health (MoH) (Zanzibar), National Bureau of Statistics (NBS), Office of the Chief Government Statistician (OCGS) and technical support from ICF international(21–24).

### Sampling technique

The survey was conducted using face-to-face questionnaire interviews and used stratified design, multistage cluster sampling to collect information about population health status, neonatal mortality, health behaviors, nutritional status family planning and demographics.

First, clusters (629) were identified and households were then selected. Among these, 26 households were systematically chosen as representative from each cluster comprising a total of 16,354 households. Eligibility for inclusion was based on all women 15-49 years old present in the sampled household the night before the interview. Detailed information on sampling procedure and design has been previously reported(12).

### Variables

The dependent variable for this study was breast cancer screening, which was measured by a question ‘Has a doctor or other healthcare provider examined your breasts to check for breast cancer?’ and the detailed information on the breast cancer screening has been published elsewhere(20). The binary response was Yes/No, following the approach employed by other researches who utilized DHS data(5,27,28). Women who were unaware of their screening status were not included in this study.

The independent variables were social-demographic and socio economic factors, furthermore we investigated the association between breast cancer screening and various factors including age, wealth index, residence, number of living children, marital status, education, health insurance, employment, pregnancy status, house hold ownership of radio or television, healthcare facility visits in the past 12 months, breast feeding status and geographical zones as reported by other researchers (5,24,27,28). We recategorized wealth index from five to three categories combining poorest and poorer as ‘poor’ middle wealth as ‘middle’ and richer and richest as ‘rich’ aligning with previous research practices(5,29–33).

Additionally, the age of survey respondents was recorded as continuous variable, was grouped into three categories 15-24, 25-34 and 35-49 years old as others researchers (5,31). Furthermore, mothers employment status was recategorized into two categories ‘working’ and ‘not working’ as previously categorized (34).

### Data management and analysis

Data cleaning and analysis were conducted using Stata 18. Categorical variables were summarized using frequencies and percentages. The Pearson Chi-squared test investigated the association between breast cancer screening and participants characteristics. Logistic regression was employed was carried out to assess associations between dependent and independent variables with 95% confidence intervals (CI). The variables associated at binary logistic regression with a significance level (p = 0.20) were entered into multiple logistic regression to identify key determinants while controlling for potential confounding effects. Statistical significance was indicated at a p-value of 0.05, and predictors of the outcome variable were identified accordingly. The variables reported were those found to be significantly associated based on adjusted odds ratios.

### Ethical consideration

The formal written request was submitted to the DHS program and approval was given to access and utilize data from http://www.dhsprogram.com. The questionnaire for standard DHS was reviewed and approved Medical Research Council of Tanzania and the Zanzibar Health Research Institute and ICF’s Internal Review Board (IRB). Participants provided either written or verbal informed consent before participating in the survey. Respondents were not subjected to any form of coercion and all data are protected ensuring no any personally identifiable information (5,25).Further details on ethical consideration are available elsewhere (26).

## Results

### Participant characteristics

Among 15,188 participants of this study, the mean (SD) age was 29.3(9.8). Almost all (94.3%) of the study participants were uninsured. Slightly less than half of the study participants were from rich households (48.7%) and the majority (64.3%) of women were from rural. Few (29.8%) of the respondents were aged between 25-34, where nearly half (48.6%) had primary education. Of all participants 92.4% were not pregnant nor did they know their pregnancy status (Table1).

The majority of women (60.1%) were either married or living with a partner. Some women in this study were not working (36.9%). Around half (52.4%) of women did not have radios in their households while some (34.2%) have television. More than half (53.8%) of the participants had history of visiting healthcare facility for the past 12 months prior the survey. Few of the study participants were breastfeeding during the survey (22.7%). Very few (5.2%) of respondents were from Southern zone and around half (53.2%) had 1-4 living children (Table1).

**Table 1:** Weighted: Self-reported breast cancer screening by participant characteristics (N=15,188)

| Variable | Total | Ever screened for breast cancer |  | P-value |
| --- | --- | --- | --- | --- |
|  | (%) | No (%) | Yes (%) |  |
| **Health insurance** |  |  |  | <0.001 |
| No | 14,302 (94.2%) | 13,664 (94.9%) | 638 (80.9%) |  |
| Yes | 887 (5.8%) | 736 (5.1%) | 151 (19.1%) |  |
| **Wealth Index status** |  |  |  | <0.001 |
| Poor | 5,015 (33.0%) | 4,929 (34.2%) | 87 (11.0%) |  |
| Middle | 2,870 (18.9%) | 2,756 (19.1%) | 114 (14.5%) |  |
| Rich | 7,304 (48.1%) | 6,716 (46.6%) | 588 (74.5%) |  |
| **Residence** |  |  |  | <0.001 |
| Urban | 5,428 (35.7%) | 4,953 (34.4%) | 475 (60.2%) |  |
| Rural | 9,761 (64.3%) | 9,447 (65.6%) | 314 (39.8%) |  |
| **Age group(years)** |  |  |  | <0.001 |
| 15-24 | 5,778 (38.0%) | 5,674 (39.4%) | 103 (13.1%) |  |
| 25-34 | 4,591 (30.2%) | 4,350 (30.2%) | 241 (30.5%) |  |
| 35-49 | 4,821 (31.7%) | 4,376 (30.4%) | 445 (56.4%) |  |
| **Education** |  |  |  | <0.001 |
| No education | 2,430 (16.0%) | 2,380 (16.5%) | 49 (6.2%) |  |
| Primary | 8,087 (53.2%) | 7,669 (53.3%) | 417 (52.9%) |  |
| Secondary/higher | 4,673 (30.8%) | 4,351 (30.2%) | 322 (40.9%) |  |
| **Pregnancy status** |  |  |  | 0.272 |
| No or unsure | 14,013 (92.3%) | 13,275 (92.2%) | 738 (93.5%) |  |
| Yes | 1,177 (7.7%) | 1,126 (7.8%) | 51 (6.5%) |  |
| **Marital status** |  |  |  | <0.001 |
| Never married | 4,022 (26.5%) | 3,910 (27.2%) | 112 (14.2%) |  |
| Married/living with partner | 9,220 (60.7%) | 8,692 (60.4%) | 528 (66.9%) |  |
| Widowed/divorced/separated | 1,948 (12.8%) | 1,799 (12.5%) | 149 (18.9%) |  |
| **Employment status** |  |  |  | <0.001 |
| Not working | 5,424 (35.7%) | 5,248 (36.4%) | 176 (22.4%) |  |
| Working | 9,765 (64.3%) | 9,153 (63.6%) | 612 (77.6%) |  |
| **Household has radio** |  |  |  | <0.001 |
| No | 7,735 (52.5%) | 7,431 (53.2%) | 303 (39.5%) |  |
| Yes | 6,998 (47.5%) | 6,533 (46.8%) | 465 (60.5%) |  |
| **Household has television** |  |  |  | <0.001 |
| No | 9,783 (66.4%) | 9,429 (67.5%) | 354 (46.0%) |  |
| Yes | 4,950 (33.6%) | 4,535 (32.5%) | 415 (54.0%) |  |
| **Visited healthcare facility last** |  |  |  |  |
| **12 months** |  |  |  | <0.001 |
| No | 7,134 (47.0%) | 6,875 (47.7%) | 260 (32.9%) |  |
| Yes | 8,055 (53.0%) | 7,526 (52.3%) | 529 (67.1%) |  |
| **Breastfeeding status** |  |  |  | 0.115 |
| No | 11,713 (77.1%) | 11,078 (76.9%) | 635 (80.5%) |  |
| Yes | 3,476 (22.9%) | 3,323 (23.1%) | 154 (19.5%) |  |
| **Zones** |  |  |  | <0.001 |
| Western zone | 1,266 (8.3%) | 1,246 (8.7%) | 21 (2.6%) |  |
| Northern zone | 1,731 (11.4%) | 1,616 (11.2%) | 115 (14.6%) |  |
| Central zone | 1,569 (10.3%) | 1,503 (10.4%) | 65 (8.3%) |  |
| Southern highlands | 924 (6.1%) | 863 (6.0%) | 61 (7.7%) |  |
| Southern zone | 803 (5.3%) | 773 (5.4%) | 30 (3.8%) |  |
| south west highlands | 1,322 (8.7%) | 1,253 (8.7%) | 69 (8.7%) |  |
| Lake zone | 4,406 (29.0%) | 4,197 (29.1%) | 209 (26.6%) |  |
| Eastern zone | 2,651 (17.5%) | 2,458 (17.1%) | 193 (24.4%) |  |
| Zanzibar | 516 (3.4%) | 490 (3.4%) | 26 (3.3%) |  |
| **Number of living children** |  |  |  | **<0.001** |
| None | 3,950 (26.0%) | 3,855 (26.8%) | 95 (12.0%) |  |
| 1-4 | 8,457 (55.7%) | 7,900 (54.9%) | 557 (70.6%) |  |
| >4 | 2,783 (18.3%) | 2,646 (18.4%) | 137 (17.4%) | **<0.001** |

### Factors associated with breast cancer screening

Logistic regression analysis was carried out to determine the association between independent variables and breast cancer screening among the study participants. Health insurance, wealth index status, residence, age group(years), education, pregnancy status, marital status, employment status, household has radio, household has television, visited healthcare facility last 12 months, breastfeeding status, zones, number of living children were found to be significantly associated with the breast cancer screening on binary logistic regression while age, wealth index, residence, education, health insurance, healthcare facility visits in the past 12 months and some of geographical zones such as northern zone, southern highlands, southwest highlands, lake zone and eastern zone on multivariate logistic regression.

Table 2 highlights the factors associated with breast cancer screening among women of reproductive age. In a multivariate logistic regression, older women aged 35 years and above were 5 times more likely to be screened for breast cancer (AOR= 5.33 95% CI 3.72, 7.63) compared to the younger participants aged 15-24. Participants with health insurance had 2 times higher odds of being screened for breast cancer compared to those without (AOR= 2.40, 95% CI 1.89, 3.06). Women from rich households had 2 times higher odds of breast cancer screening compared to those from poor (AOR= 2.34, 95% CI 1.61, 3.38). Women residing in rural areas were 41% less likely to be screened for breast cancer compared to those in urban areas (AOR= 0.59, 95% CI 0.46, 0.763).

**Table 2:**
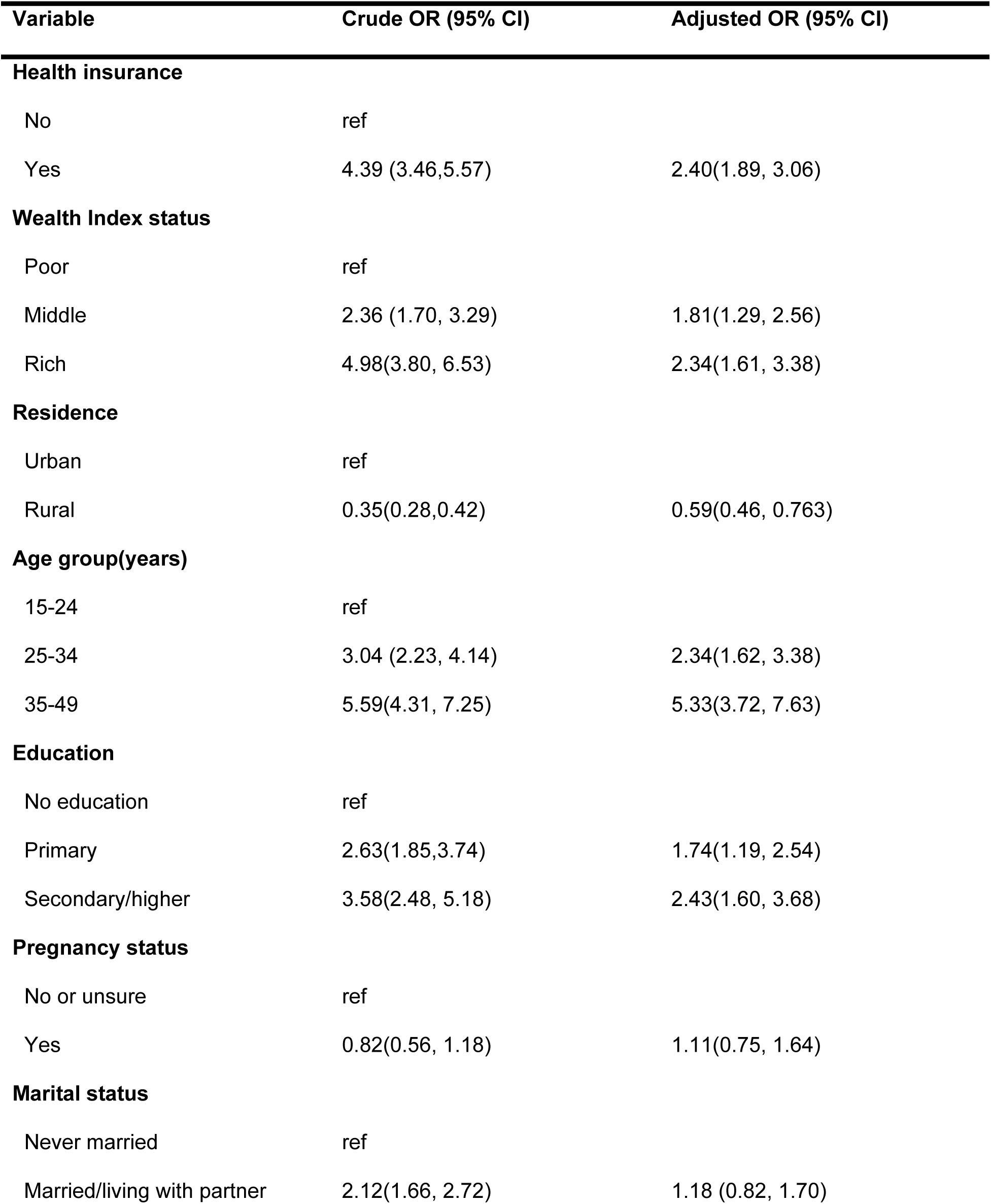
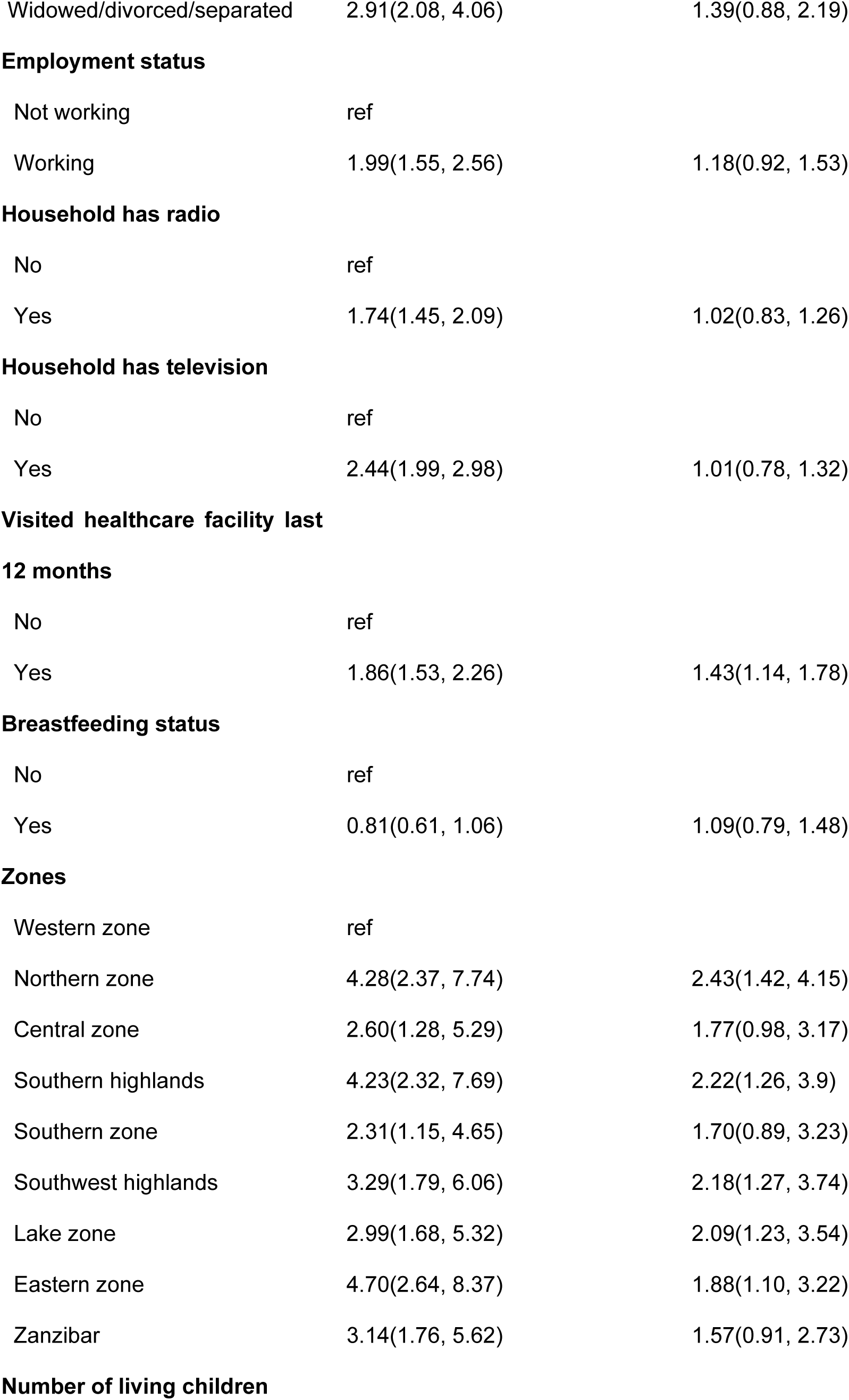
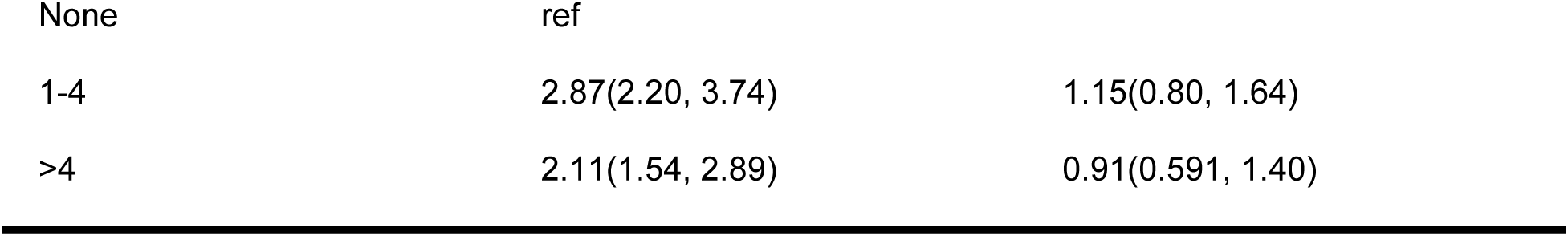
Factors associated with breast cancer screening among women of reproductive age in Tanzania.

## Discussion

This cross-sectional study assessed factors associated with breast cancer screening among women of reproductive age in Tanzania, A sizable sample of women participated in this survey, which helped us identify screening drivers and offers insights into their background characteristics, after controlling for other determinants the following factors remained significant predictors for breast cancer screening; age, wealth index, residence, education, health insurance, healthcare facility visits in the past 12 months and some of geographical zones such as northern zone, southern highlands, southwest highlands, lake zone and eastern zone.

The occurrence and fatality rates of breast cancer have experienced a rapid rise in developing nations like Tanzania and a decline in developed countries(2).Radiologist, Oncologist, breast surgeons and pathologists who play a crucial role in early detection and planning of the treatment are still scarce in Tanzania(6).

Age emerged as a very influential predictor, this could be explained by older women are vulnerable and are more knowledgeable about breast cancer screening(9,13,14).This findings are consistent with previous studies which also found older women were more likely to screen for breast cancer than younger women(5,27). Most of the health care facilities recommend women aged 40 years and above to undergo breast cancer screening to prevent them from breast cancer(35). On the other hand, most SSA countries do not have national screening programs as well as enough funds to screen all eligible women(35).

In Tanzania particularly, there is unclear and unstandardized protocol for early breast cancer screening on top of that inefficient referral system adds cost and delay most of the clients with breast lesions(11). Due to poor financial and human resources in SSA, yearly clinical breast examination to women under 40 years may be useful way to find early signs of breast cancer (36–39). Previous researches have shown that 20% of breast cancer occur among women aged 30-40 years (37)(40,41). Moreover, a study indicated that in SSA only 2.2% of women aged 40-69 years screened for breast cancer in the past 5 years (42). Furthermore, a study done in Sudan depicted that engaging local community volunteers for clinical breast examination might potentially enhance the early diagnosis of breast cancer in women who do not exhibit any clinical symptom(43).

As one might anticipate, and in line with previous studies(5,27,28), there was a positive association between health insurance coverage and breast cancer screening. This may be the case due to the fact that having health insurance gives women the chance to receive preventative treatment at no additional expense. We found that wealth was positively associated with breast cancer screening, wealthy women are better able to afford health insurance which result in receiving preventive care services with no or minimum cost, on the other hand poor women are less likely to prioritize preventive care services over their daily needs (5,42).

The negative association between breast cancer screening and living in the rural was not unexpected and can be explained by uneven access to healthcare services and insufficiency of health care facilities that can offer breast cancer screening compared to urban(5,28,44).

Educational attainment demonstrated a positive association with screening, educated women may know the harmful effect of breast cancer and early detection measures (45,46) and this showcase the role of education in promoting health-seeking behaviors, our study is in accordance with earlier studies which found that educated women were more likely to be screened for breast cancer than those with no education(27,28).

Notably, healthcare facility visits in the past year was positively associated with breast cancer screening, this emphasizes the impact of regular health check-ups and it aligns with the findings from previous study which found that women who visited healthcare facility in the past 12 months were more likely to screen for breast cancer compared to those who didn’t (5).

Unlike other studies(5,27), our study found that mass media exposure(having television or radio in the household) was not associated with breast cancer screening after controlling for other factors this can be explained by lack of political will on promoting breast cancer screening as well as logistical challenges in accessing screening services, cultural considerations and socioeconomic disparities among Tanzania women of reproductive age.

### Strength and limitation

This is population-based cross-sectional study of more than 15, 000 women of reproductive age in Tanzania, the findings may contribute to improving breast cancer screening uptake among women of reproductive age. The limitation of this study include, the study was limited to only women of reproductive age 15-49 and evidence suggest that median age of breast cancer diagnosis is 62 (47).Also, the study is prone to recall bias because the response was self-reported, additionally DHS does not capture timing of the breast cancer screening which is essential for early diagnosis and treatment. Lastly, the study is a cross sectional nature of the survey which does not allow for the determination of temporal relationships

### Conclusion and recommendation

Very few women of reproductive age in Tanzania get screened for breast cancer, it is crucial to address this escalating burden of breast cancer through heightened health awareness, effective prevention strategies, and enhanced access to medical treatment. Older age, being wealthy, residing in rural areas, being educated, having health insurance, healthcare facility visits in the past 12 months and some of geographical zones such as northern zone, southern highlands, south west highlands, lake zone and eastern zone were independently associated with breast cancer screening. These findings suggest that focused interventions are needed to lower the disease’s incidence and increase survivor rates. Women of reproductive age should be made aware of the advantages of breast cancer screening through extensive health education and awareness programs. Making breast cancer screening easier to afford and reach, especially in rural regions, is vital. Emphasizing and executing Universal Health Insurance (UHI) could help more financially constrained women access breast cancer screenings. Future studies should make use of qualitative and longitudinal approaches so as to explore deeply as per why women of reproductive age don’t screen for breast cancer. Data regarding the time of breast cancer screenings should be gathered for future surveys conducted in Tanzania. The survey should be improved in order to precisely record the particular kind of screening that was carried out, such as mammography or clinical breast examination (CBE).

## Funding

This research did not receive any specific grant from funding agencies in the public, commercial, or not-for-profit sectors.

## Authorship contributions

**Jovin R Tibenderana:** Conceptualization, Data curation, Formal analysis, Investigation, Methodology, Project administration, Supervision, Validation, Visualization, Writing – original draft, Writing – review & editing. **Sanun Ally Kessy:** Investigation, Data curation, Writing – review & editing, Visualization. **Dosanto Felix Mlaponi:** Investigation, Data curation, Writing – review & editing, Visualization. **Ndinagwe Lloyd Mwaitete:** Investigation, Data curation, Writing – review & editing, Visualization. **John Elyas Mtenga:** Investigation, Data curation, Writing – review & editing, Visualization, Supervision, Validation.

## Disclosure

The authors report no conflicts of interest in this work.

## Abbreviations

AOR: Adjusted odds ratio
CI: Confidence Interval
COR: Crude odds ratio
DHS: Demographic Health Survey

## Data Availability

Data are not owned by the authors. Data are available from Tanzania Demographic and Health Surveys (DHS) program (https://dhsprogram.com/Data/). DHS data may be accessed through registration of a research project on the DHS website and requesting access to data. This is the same manner the authors obtained access to the data. The authors did not have any special access privileges to DHS data

## Notes

### Competing Interest Statement

The authors have declared no competing interest.

### Funding Statement

The author(s) received no specific funding for this work.

### Author Declarations

The formal written request was submitted to the DHS program and approval was given to access and utilize data from http://www.dhsprogram.com. The questionnaire for standard DHS was reviewed and approved Medical Research Council of Tanzania and the Zanzibar Health Research Institute and ICF’s Internal Review Board (IRB). Participants provided either written or verbal informed consent before participating in the survey.

